# Analysis of the outbreak of COVID-19 in Japan on the basis of an SIQR model

**DOI:** 10.1101/2020.06.02.20117341

**Authors:** Takashi Odagaki

## Abstract

The SIR model is modified, which may be called an SIQR model, so as to be appropriate for COVID-19 which has the following characteristics: [1] a long incubation period, [2] transmission of the virus by asymptomatic patients and [3] quarantine of patients identified through PCR testing. It is assumed that the society consists of four compartments; susceptibles (S), infecteds at large (simply called infecteds) (I), quarantined patients (Q) and recovered individuals (R), and the time evolution of the pandemic is described by a set of ordinary differential equations. It is shown that the quarantine rate can be determined from the time dependence of the daily confirmed new cases, from which the number of the infecteds at large can be estimated. The number of daily confirmed new cases is shown to be proportional to the number of infecteds a characteristic time earlier, and the infection rate and quarantine rate are determined for the period from mid-February to mid-April in Japan, and transmission characteristics of the initial stages of the outbreak in Japan are analyzed. The effectiveness of different measures is discussed for controlling the outbreak and it is shown that identifying patients through PCR testing and isolating them in a quarantine is more effective than lockdown measures aimed at inhibiting social interactions of the general population. An effective reproduction number for infecteds at large is introduced which is appropriate to epidemics controlled by quarantine measures.

## Introduction

The pandemic (COVID-19) due to the novel coronavirus (SARS-CoV-2) is still expanding throughout the world 6 months after the first outbreak in November 2019 in China (*1*). Compared to other viral outbreaks such as Influenza, for which the SIR model is applied, COVID19 has the following characteristics (*2, 3*). [1] A long incubation period. [2] Prevalence of asymptomatic transmission. [3] Detection of infection by PCR tests. The biggest challenge in modeling the COVID-19 outbreak is that a large number of patients are hidden in the general population as they continuously infect others, and that the only observable metric on which we can base estimates is the number of daily new cases confirmed by positive tests.

Traditionally, the dynamics of infectious diseases have been understood by the deterministic compartmental models described by differential equations such as the SIR model (*4*) and the SEIR model (*5, 6*) or by the stochastic model described by a renewal integral equation (*7*). In the SEIR model, the population consists of susceptibles (S), exposed individuals (E), infectious individuals (I) and removed individuals (R). The SIR model does not include the compartment of exposed individuals. In these compartmental models, quarantine is considered to be an efficient way for medical treatment of patients and one of the removal processes. The removal time is usually treated as the scale of time, and thus the quarantine rate is included only in the reproduction number. The SIR-X model proposed by Maier and Brockmann (*8*) includes the quarantine process in the analysis of the early state of the outbreak in China.

In the renewal model, the number of infecteds at a given time is assumed to be determined by a convolution of a time-dependent transmission coefficient and the number of infecteds at earlier times with the reproduction number as a multiplier, and the quarantine process is hidden in the reproduction number.

There are several attempts to understand the nature of the outbreak of COVID-19 on the basis of the SEIR model (*8,9*) and the renewal model (*10*). Apparently no attempts have treated quarantined patients as a compartment and the quarantine rate as a controllable parameter, although it plays an essential role in the measures against the COVID-19. At this moment in the middle of outbreak of COVID-19, it is important to establish a universal, predictable and simple model for the outbreak which can be understood by citizens and policy makers.

In this paper, keeping the characteristics of the COVID-19 explained above in mind, I introduce four compartments; susceptibles (S), infecteds at large or simply infecteds (I), quarantined patients (Q) and recovered individuals (R), and describe the transmission process by a system of ordinary differential equations where the quarantine rate of infecteds is introduced as a key parameter. This model may be called an SIQR model. Although the basic differential equations are similar to those of the SIR model, the present SIQR model treats the quarantine process as a measure controlled by policy. Namely, I consider the quarantined patients not only as individuals to be treated in the hospital but also as key players in combating the spread of the virus. Figure 1 shows the flow diagram of the SIQR model.

I first show that the number of infecteds at large can be estimated from the daily confirmed new cases. Then, I present possible scinarios of the outbreak, considering three hypothetical countries. I analyze the time dependence of the daily confirmed new cases reported in Japan till the end of April which is shown in Fig, 2. Using the solution of the SIQR model, I fit the data from March 1st to April 29th by four pieacewise exponential functions, and compare the time evolution of parameters derived from fitting with the measures taken by the Japanese government from mid-February to mid-April. This period saw a rapid rise in daily confirmed new cases, and is also notable for a succession of policy milestones, one being the planned Olympiad postponed by IOC on March 24th, and the other being the prime minister announcing on April 5th that a state of emergency would be formally declared two days later on the 7th. The government issued stay-at-home request, asking 80% reduction of social contact. I compare the time dependence of daily confirmed new cases if different measures would have been taken at the beginning of April in Japan. I compare various policies characterized by a combination of differing degrees of testing/quarantine and societal lockdown and show that the quarantine measure is more efficient than the lockdown measure.

**Figure 1:**
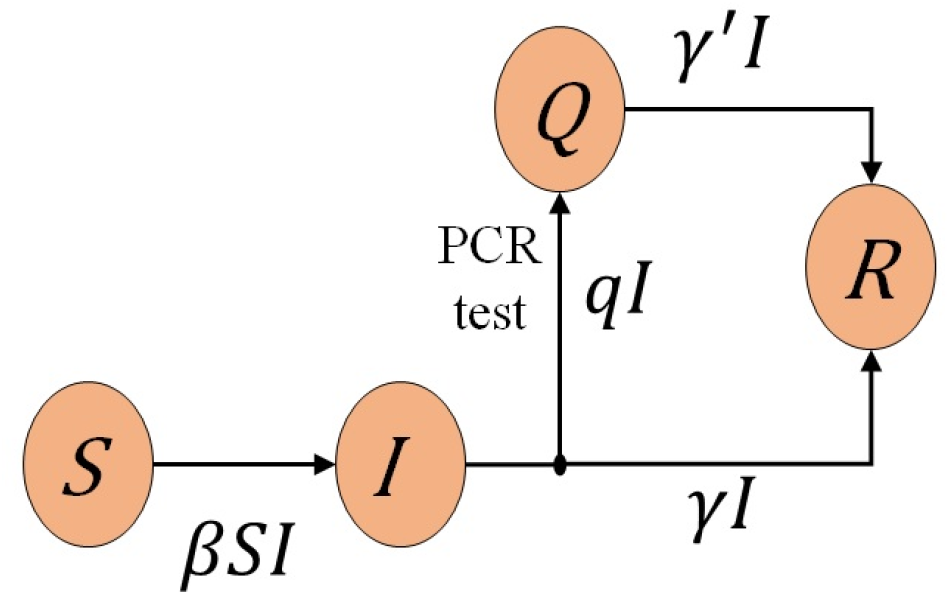
Flow diagram of the SIQR model. If the quarantine process is included in the recovery process and *γ* = *γ*′, is assumed, the SIQR model is reduced to the SIR model.

**Figure 2:**
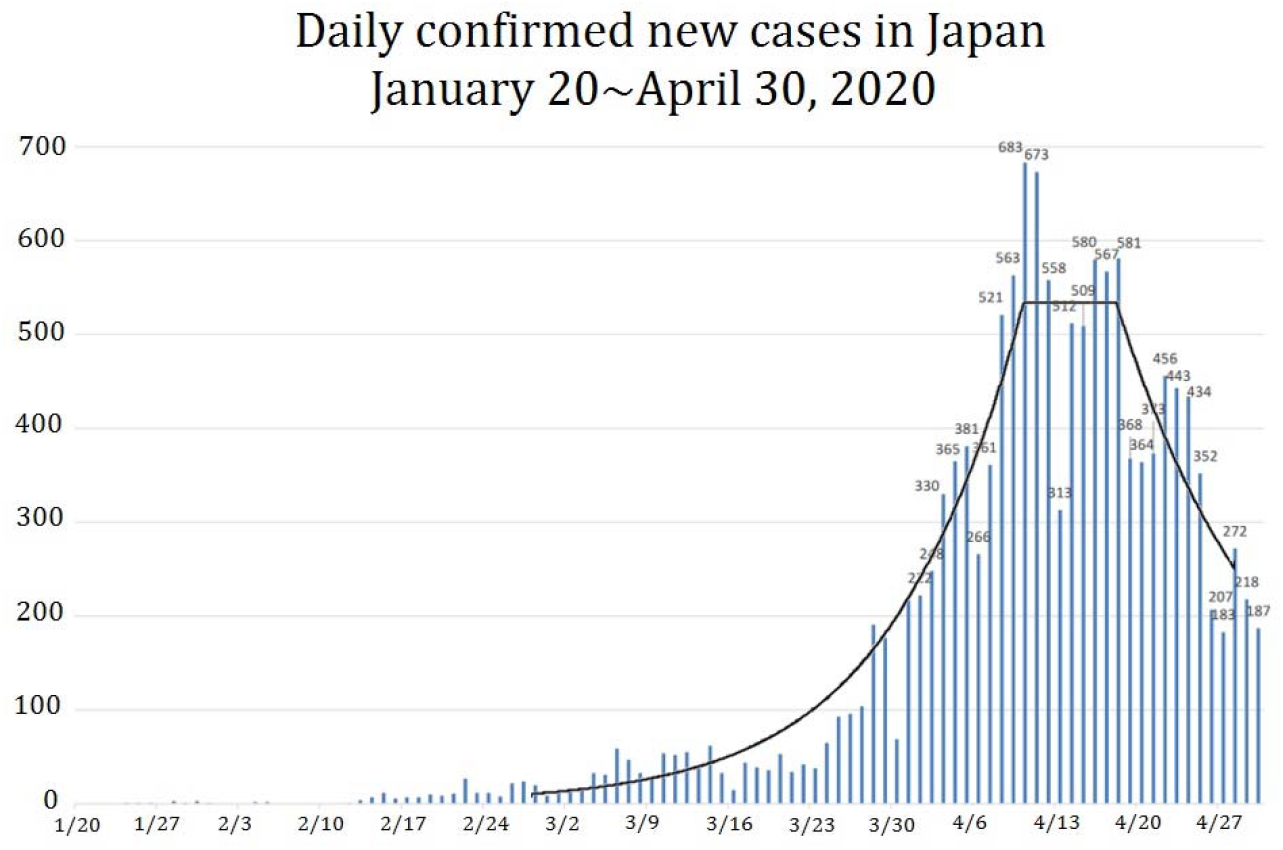
The daily confirmed new cases in Japan till the end of April. The solid curve represents fitting by four piecewise exponential functions with parameters and periods given in Table 2. Data are from Ministry of Health, Labor and Welfare, Japan (*11*).

## SIQR model

The dynamics of the SIQR model are described by the following set of ordinary differential equations:

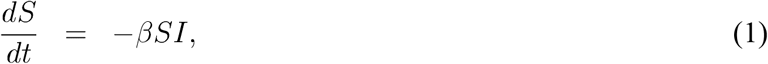

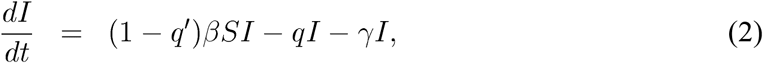

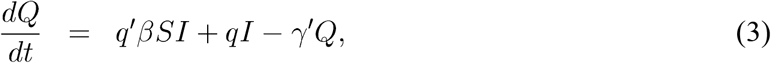

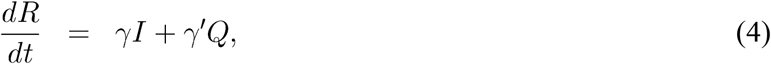

where *t* is the time, and *S*, *I*, *Q* and *R* are the stock of susceptible population, infecteds at large, quarantined patients and recovered (and died) patients, respectively. Here, I assumed that the net rate at which infections spread is in proportion to the number of encounters between susceptibles and infecteds at large, *βSI*, where *β* is a transmission coefficient. The parameter *q*′ represents a per capita rate of quarantine of the new patients immediately after they get infected. For COVID-19, *q*′ can be set to zero since the incubation period is long. Infecteds at large, regardless of wheter they are symptomatic or asymptomatic, are quarantined at a per capita rate *q* and become non-infectious to the population. Quarantined patients recover at a per capita rate *γ*′ where 1/*γ*′,is the average time it takes for recovery) and infecteds at large become non-infectious at a per capita rate *γ* where 1/*γ* is the average time that an infected patient at large is capable of infecting others). It is apparent that Eqs. (1) ~ (4) guarantee the conservation of population *N = S + I + Q + R*.

If we assume *γ* = *γ*′ and regard *Q* + *R* as the recovered compartment, Eqs. (1) ~ (4) are identical to those of the SIR model with removal rate of infecteds *q* + *γ*. Since the value of *q* depends strongly on government policies and the only observable is the number of quarantined patients on each day, the SIQR model which treats the quarantine process explicitly is superior to the SIR and SEIR models in the analysis of the outbreak of the COVID-19.

In the following discussion, I concentrate on the number of infecteds in the early stage of the outbreak where *I* + *Q* + *R* << *N* is satisfied so that I can assume *S* ≃ *N*. Then the basic equations of the SIQR model become

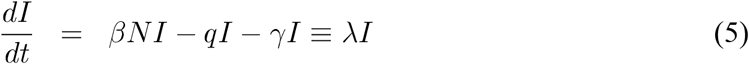

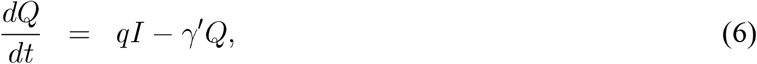

where the rate of change of the infecteds

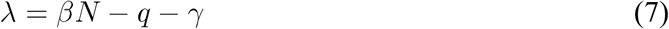

determines the short-term behavior of the number of infecteds.

The parameter *β* determines the initial growth of infecteds and is determined by the cumulative effect of the virulence of the virus and social factors. The longer latent period and shorter infectious periods reduces the number of infectious patients in the infecteds at a given time which in turn makes *β* effectively smaller. Similarly, the shorter incubation period and the higher onset probability increases the number of quarantined individuals which makes the quarantine rate *q* larger.

The parameters are dependent on the lifestyle of each community and measures taken by governments. For example, availability of a vaccine, implementation of social distancing or lock down measures will all decrease *β*, while widespread PCR testing and isolation of patients will increase *q*. Although the recovery rate for quarantined patients *γ′*, will be increased when a new drug for SARS-CoV-2 is developed, it would not increase *γ* since a patient is unlikely to seek treatment unless they are first identified.

It should be noted that the characteristics of the outbreak of the COVID-19 can be understood by effective values of three parameters *β*, *q* and *γ*,

It is usual to take the inverse of the removal rate (*q* + *γ*) or the recovery rate (*γ′*) as the scale of time. However, these parameters are policy dependent and cannot be considered as an intrinsic time scale of the problem. In order to present results in the style that citizens and policy makers can easily understand, I take one day as the scale of time in this paper.

Although an exact solution to Eqs. (1) ~ (4) can be obtained by introducing a scaled time when parameters do not depend on time (*12*), the solution is impractical for the purpose of analyzing the outbreak and predicting the outcome of various measures.

## Estimation of the number of infecteds at large

The term *qI*(*t*)in Eqs. (5) and (6) is the number of daily confirmed new cases Δ*Q*(*t*)and thus the number of infecteds is given by

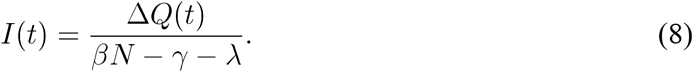

This relation indicates that if we can determine λ from the time dependence of the daily confirmed new cases, *β* from the initial increase of infecteds and infectious period *γ*^−1^ of asymptomatic patients, we can determine the number of infected at large from the daily confirmed new cases.

In the data reported in Japan (*11*), the daily confirmed new cases are apparently a constant around ~ 530 in the period from 11th to 19th of April, 2020 as shown in Fig. 2. If this is the case, λ must be negligibly small in this period, and thus

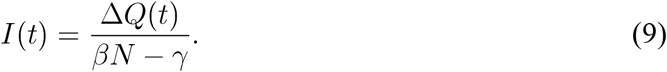

As will be seen later, λ ≃ 0 and *q* ≃ 0.096 in this period when *γ* = 003 (or 33 days of the latent plus infectious periods) is used, and thus I find

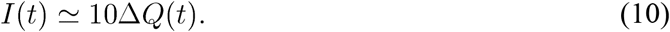

This means that there were about 5300 infecteds in Japan at the beginning of April. This number increases to 8030 if *γ* = 006 (17days for the recovering time) is used.

## Time evolution of outbreak

In the time period when the parameters do not depend on time, Eqs. (5) and (6) can be readily solved:

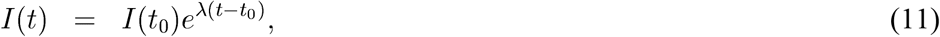

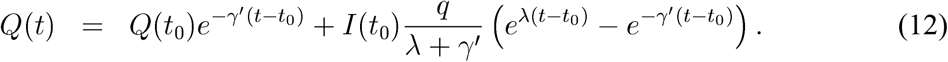

Equation (11) indicates that the number of infecteds is determined by λ and its initial condition. The parameter λ depends on measures that the government employs, and we can assess the measures from the time dependence of the number of infecteds.

At the beginning of the outbreak, infecteds cannot be detected since symptoms do not appear for the incubation period and thus cannot be quarantined. Therefore, the number of infecteds increases exponentially as

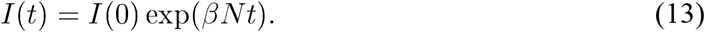

About a few weeks later *t*_1_ (*13*), some infecteds will recover and thus the growth rate will be reduced by *γ*:

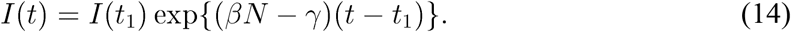

After some time, a certain number of patients with severe symptoms will appear and the government will introduce various measures at time *t*_2_ such as social distancing, and societal lockdown measures. If the measures in which *β* is reduced to (1−*x*) *β* keeping *q* as before is effective, the number of infecteds will start to decline exponentially, following

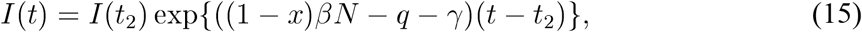

where *x* represents the degree of social distancing and lockdown. The effectiveness of the measure can be judged by the size of the decay constant |((1 − *x*) *βN* − *q* − *γ*)|.

If no measures are taken, the number of infecteds increases exponentially at the beginning. This increase is limited by the nonlinear term *βSI* in Eqs. (1) ~ (3), which is neglected in the present study concentrating on the early stage of the outbreak. The nonlinear term makes the infection trajectory falling off after reaching a maximum. Various measures taken by the government appear as abrupt changes in the infection trajectory.

In order to see the effects of measures, I consider three different model countries, U, J and K. I assume when no measure is taken, the doubling time of infecteds is 7 days and the recovering time is 30 days, that is *βN* = 0.099 and *γ* = 0033.

For the first 30 days, the number of infecteds simply increases exponentially with λ = 0.099, and for the next 30 days it increases with reduced rate λ = 0.066 due to the recovering process. In the following two periods, days 60 ~ 75 and days 75~, each country takes different measures. Country U introduces a strict PCR testings and qurantine *q* = 0.001 in the first period and in the second period 50% reduction of social contact *x* = 05and a strong quarantine measure *q* = 0.1. Country J introduces a moderate quarantine measure *q* = 0.066 in the first period. In the second period, stay-at-home request with 80% reduction of outings is issued, keeping the same quarantine measure. Country K introduces a complete lockdown preventing all social contact *x* = 1 and strong quarantine measure *q* = 0.1 from the first period. Table 1 summarizes the parameters of each model country.

**Table 1:** Parameters for three hypothetical model countries, U, J and K. λ = (1 − *x*) *βN* − *q* − *γ* where *βN* = 0.099 and *γ* = 0.033. *x* represents the degree of societal lockdown and *q* depends on the quarantine measure of each country.

|  | days(60~75) |  |  | days(75~) |  |  |
| --- | --- | --- | --- | --- | --- | --- |
| | $x$ | $q$ | $\lambda$ | $x$ | $q$ | $\lambda$ |
| U | 0 | 0.001 | 0.065 | 0.5 | 0.1 | -0.083 |
| J | 0 | 0.066 | 0 | 0.8 | 0.066 | -0.079 |
| K | 1 | 0.1 | -0.133 | 1 | 0.1 | -0.133 |

Figure 3 compares the time dependence of infecteds for the three hypothetical model countries.

Next, I will take a look at the relation of *I*(*t*) to the number of daily confirmed new cases Δ*Q*(*t*). I assume that patients tested positive are quarantined either in a hospital or at home, and are no more infectious to the population. The daily confirmed new cases Δ*Q*(*t*) is proportional to the number of infecteds who have an onset of symptoms. Since the latter is given by a convolution of the incubation period distribution function *ψ*(*t*) and the number of infecteds *I*(*t*). Therefore, Δ*Q*(*t*) can be expressed as

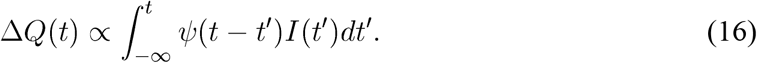

**Figure 3:**
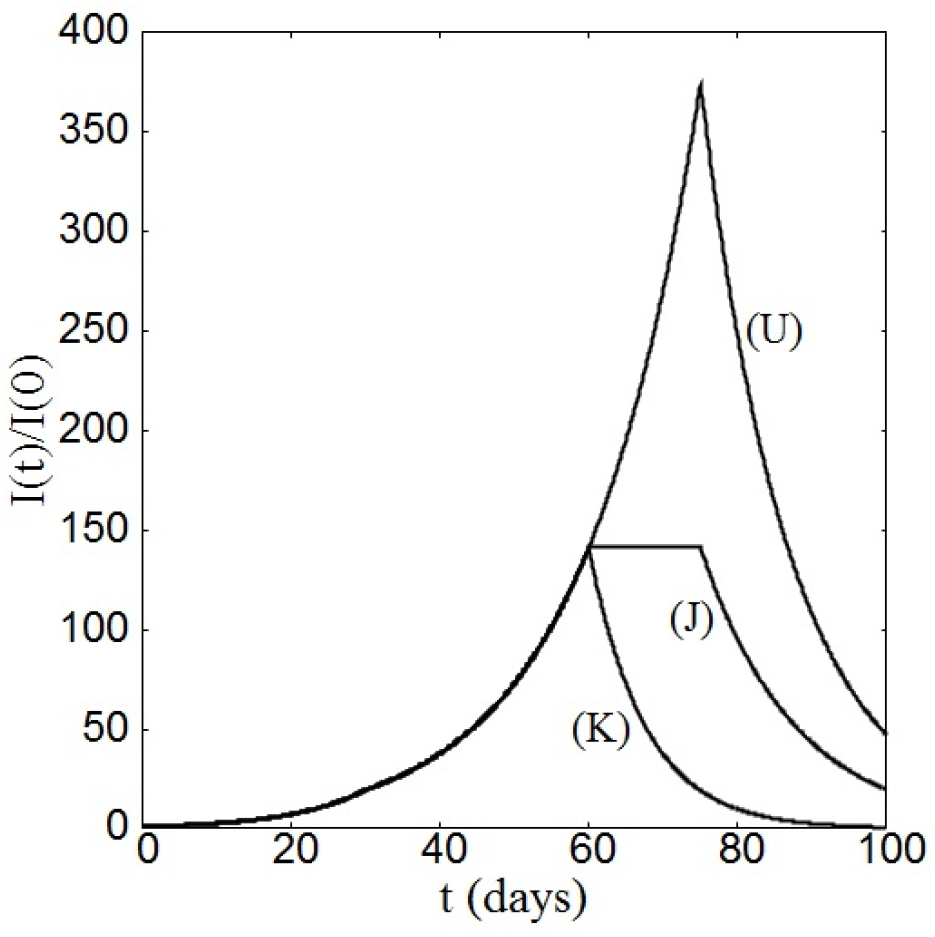
Comparison of three model countries. For the first 60 days, no difference is set. See text and Table 1 for the detailed setting of the parameters for the latter half.

The incubation period distribution is a well behaved function with a single peak (*14, 15*). I assume, for the sake of simplicity, that *ψ*(*t*) is given by

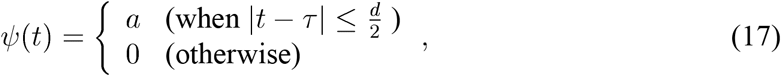

where *a* is a positive constant and *d* denotes the incubation period. When *I*(*t*) = *I*(0) exp (*λt*), it is straitforward to show that (*16*)

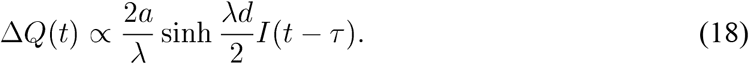

Therefore, the daily confirmed new cases is related to the number of infecteds mean-incubation-time earlier. In fact, the number of daily confirmed new cases of many countries seem to be represented by piecewise exponential functions (*17*).

I analyze the daily confirmed new cases Δ*Q*(*t*) in Japan by fitting it with piecewise exponential functions, where I neglected the effect of the transition periods. From the fitting, I determined the rate of change λ, from which parameters *βN* and *q* are estimated. The solid curve in Fig. 2 shows a piecewise fitting in the period of March 1st ~ April 29th by four exponential functions with λ listed in Table 2.

**Table 2:** Parameter λ used for fitting in Fig. 2 and estimated values of other parameters. Numbers in (⋯) are estimated from other data and numbers in [⋯] are inherited from the previous period. γ is fixed in the entire period.

| Period<br>(14 days earlier) | 3/1∼4/2<br>(2/16∼3/19) | 4/2∼4/11<br>(3/19∼(3/28) | 4.11∼4/19<br>(3/28∼4/5) | 4.19∼ 4/29<br>(4/5∼4/15) |
| --- | --- | --- | --- | --- |
| $\lambda$ | 0.096 | 0.090 | 0.0 | $-0.075$ |
| $\beta N$ | 0.126 | [0.126] | [0.126] | 0.051 |
| $q$ | (0) | 0.006 | 0.096 | [0.096] |
| $\gamma$ | (0.03) | (0.03) | (0.03) | (0.03) |

From the λ value determined by the fitting, other parameters can be estimated. I assume that parameters do not change unless effective measures are introduced. First I set γ = 0.03 in the entire period assuming that the recovery time of infected at large is 33 days. Here I took a longer period among widely distributed observations (*13*). Next, I set *q* = 0 in the first period since virtually no infecteds were quarantined in this period. The estimated values for *βN* and *q* are listed in Table 2.

Several comments are in order: (1) the basic reproduction number *R*_0_ *β*/(*q* + γ)is estimated to be ~4 for the first two periods and becomes 0.4 after April 5th passing the period where *R*_0_ ~ 1. (2) The reduction of *βN* after April 5th is about 0.051/0.126×100 = 40% which may be attributed to the emergency declaration by the government on April 7th. Apparently the effect is seen two days earlier since the government announced it on April 5th in advance. (3) The effectiveness of PCR test and quarantine, that is the probability of a patient being qurantined when symptoms appear, is rather low around 25 ~ 50% due to the strict conditions set by the Japanese government for PCR tests (such as a mandatory 4 day waiting period before a symptomatic patient is allowed a test). Since *q* ~ onset rate × the probability of getting PCR test, the onset probability is estimated to be 20 ~ 40%. These values are in the wide range 20 ~ 95% of estimations by other researchers (*18*). (4) Since the fitting is very much rough as the data are, one might fit the data by different sets of exponential functions, and the estimated value of parameters might be a little different from those obtained above.

## Comparison of possible measures

If the estimation given in the previous section is correct, there must have been 5300 infecteds at large in Japan at the beginning of April. In this section, I compare how the number of daily confirmed new cases Δ*Q*(*t*)would have reduced if Japanese government had taken different measures.

A measure can be characterized by two parameters *x* and *y*, which define an effective decay constant λ(*x*, *y*) as

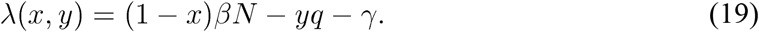

For the purposes of this paper, I will treat social distancing and lockdown measures as part of the same effort to reduce social contact, whose cumulative effectiveness is represented by *x* (0 ≤ *x* ≤ 1), with *x* = 0representing no change to social behavior, and *x* = 1 representing the most extreme form of societal lockdown.

Parameter *y* denotes the enhancement factor of quarantine rate. Since (1− *x*) *βN* − *yq* − γ is supposed to be negative when the number of infecteds is declining, the smaller (1 − *x*) and the larger yare, the decay rate |λ(*x*, *y*)| becomes larger.

From the values of *βN* in the third and fourth periods in Table 2, the reduction rate of social contact is estimated to be 60% after the emergency declaration. Therefore, I compare, relative to the third period of Table 2 (*βN* = 0.126, *q* = 0.096, *x* = 0 and *y* = 1), hypothetical measures of (1) 80% reduction of outings that the government requested and at the same level of the PCR test (*x* = 0.8, *y* = 1) and (2) 100% reduction of outings, that is the strictest form of lockdown, and at the same level of the PCR test (*x* = 1.0, *y* = 1). I also estimate the reduction rate for (3) 50% reduction of outings and doubled quarantine rate by introducing an expanded policy for PCR testing (*x* = 1.5, *y* = 2) and (4) quadrupled quarantine rate without any restriction on social interactions (*x* = 0.0, *y* = 4). Figure 4 compares the reduction rate of the daily confirmed new cases for different measures, had they been taken from April 5th. Figure 4 shows clearly that partial or strict lockdowns are less effective than increase of the quarantine rate. Countries like Korea, Vietnam and Taiwan which took strong lockdown and quarantine measures have apparently succeeded in controlling the first wave outbreak of COVID-19 (*19-21)*.

**Figure 4:**
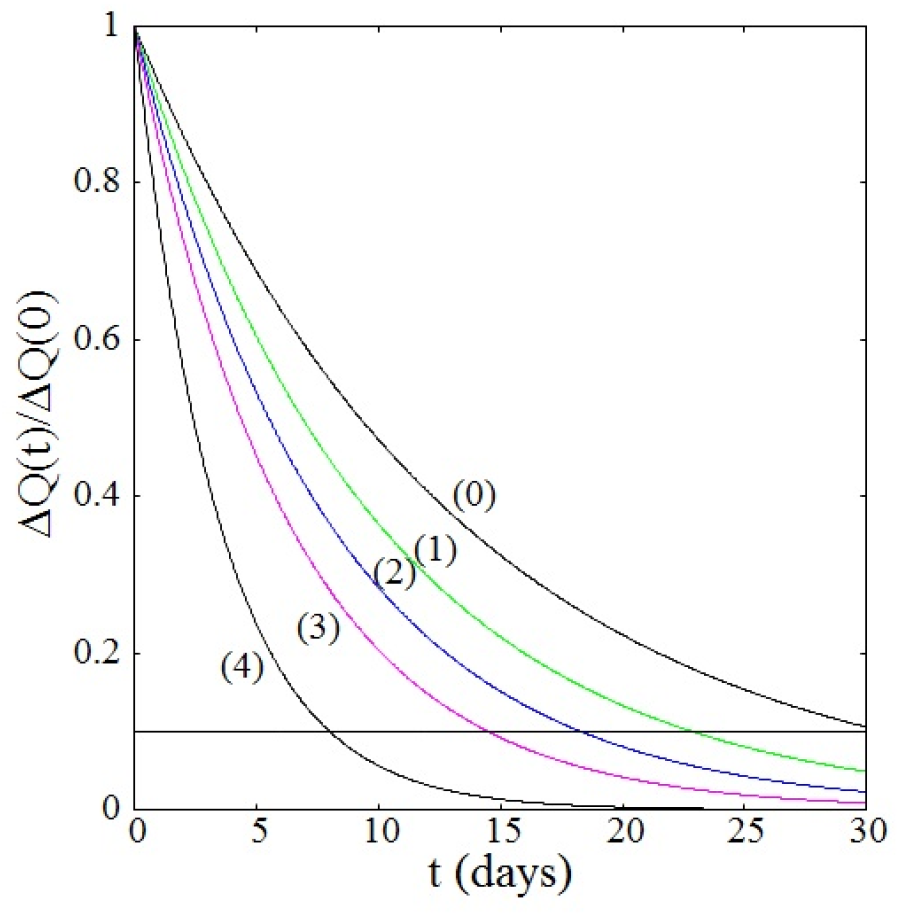
Comparison of reduction rate of the daily confirmed new cases for four different hypothetical measures, relative to the third period of Table 2, if it was taken on April 5th (*t* = 0). Observed decay given in the fourth period is also shown by curve (0). If *γ*, is set to a larger value, the effect in decay rate due to the enhanced quarantine rate becomes somewhat moderate. The horizontal line represents Δ*Q*(*t*)/Δ*Q*(0) = 0.1.

In order to see the effectiveness of measures characterized by parameters *x* and *y*, the effective decay rate |λ(*x*, *y)|* is plotted as a function of *x* and *y* in Fig. 5. The decay constant Eq. (19) shows that *y* has more leverage than *x* in making ((1 − *x)βN − yq −* γ) a large negative value. Therefore, we see that increasing the quarantine rate is a more effective measure than any forms of societal lockdown. Although this figure depends on the value assumed for the parameters, effectiveness of the quarantine does not change (*22)*.

**Figure 5:**
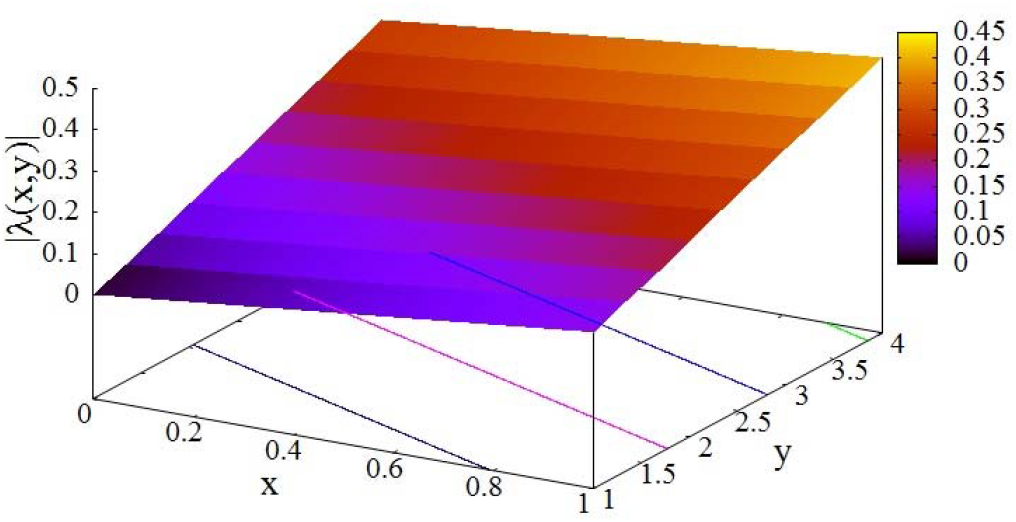
Dependence of the effective decay rate |λ(*x*, *y)|* on parameters *x* and *y* which represent the strength of lockdown and the enhancement factor of the quarantine rate, respectively. The values for *βN*, *q* and γ are taken from the third period of Table 2.

## Discussion

The pandemic COVID-19 has unusual nature such as infectious asymptomatic patients and a long incubation period in comparison to known epidemics (*1*). In the typical epidemics, all infected individuals become infectious after a latent period and show onset of symptoms after an incubation period. Therefore the number of infecteds can be estimated from the number of symptomatic patients. We cannot do the same for COVID-19, however, because of the prevalence of asymptomatic patients in the general population. Short of testing the entire population in a single day, the only visible metric we have for the purpose of estimating the total number of infecteds is the number of daily confirmed new cases Δ*Q*(*t)*. Therefore, what we need is to construct a physical model for the pandemic in which Δ*Q*(*t)* is treated explicitly and can be related to the number of infecteds at large *I*(*t*).

The infection process itself is a complex phenomenon with numerous parameters. What we need now is a simple theory in which those parameters can be effectively absorbed in essential parameters describing COVID-19. I have presented in this paper a universal, predictable and simple model which is constructed on the basis of the mean field theory and distinguishes the quarantine process explicitly. In order to determine parameters of the model, I fitted the data of the daily confirmed new cases reported in Japan. Using the time dependence of the parameters, I analyzed the characteristics of the outbreak in Japan from mid-February to mid-April. Because of the advance notice of the declaration of the national emergency expected on April 7th, the effect is seen apparently from April 5th. After April 5th, the partial lockdown appears to have resulted in 60% reduction of the social contacts, which is 20% less than the value 80% that the government requested, Has the government introduced other measures as well at the same time, the reduction of infecteds would have been much faster. In particular, the four-day waiting period required for symptomatic patients before they were allowed a PCR test has made the situation much worse. If the waiting period was halved, the quarantine rate would have been doubled. The curves (3) and (4) in Fig. 4 clearly show the effect of efficiency of the PCR test. The Japanese government could have taken much better and stronger measures.

The basic equation can be modified so as to include the memory effect in the infection process and the quarantine process. To this end, the first two terms in the middle part on the right-hand side of Eq. (5) can be written as

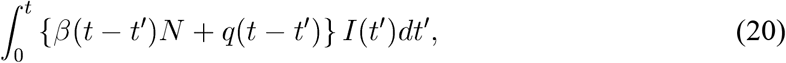

where *β*(*t − t′)* denotes the time dependence of the transmission coefficient and *q*(*t − t′)* represents the rate of quarantine of infecteds at time *t* who got infected at time *t*′,

In order to check the consistency of the present analysis in Japan, I verified that the daily confirmed new cases in Japan up until the first two weeks of May are on the curve (0) in Fig. 4.

Although the present model is based on the same compartmental approach as the SIR and SEIR models, it proposes a novel concept in that the quarantine is an effective measure in combatting the spread of the COVID-19 (*22*). According to the SIR model apparently employed by the Japanese government, the quarantine was considered as a part of treatment and the last two terms in Eq. (2) *qI + γI* are considered together for effective treatments in a hospital. Therefore, many doctors in Japan were strongly against increasing the number of PCR tests, fearing the collapse of the medical care system and false positives in PCR tests leading to placing healthy individuals in quarantine. The policy on quarantine set by the Japanese government was not appropriate to COVID-19.

Since the early stages of civilized history, the quaratine process was considered an effective method for reducing the spread of infection. Therefore, the term (*βN* − *q)I* in Eq. (5) is considered the number of infecteds newly added in the population, and the quarantined *−pI* is considered a means of removing infecteds from the population. It is apparent that increasing the quarantine rate is more effective in making λ(*x*, *y*) negative than lockdown measure, since the former can reduce λ(*x*, *y*) by several times of *q* while the latter can contribute at most *βN*.

For COVID-19, we should go back to this tradition with contemporary method in identifying pre-symptomatic and asymptomatic patients by PCR tests. In this sense, an effective reproduction number *R_e_* relevant to COVID-19 should be defined by

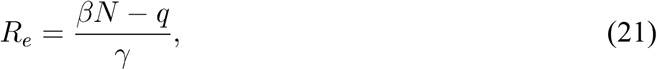

which represents the rate of change of infecteds at large. The effective reproduction number in Japan in each period shown in Table 2 is 4.2, 4.0, 1 and -1.5, where *R_e_* = 1 indicates that the number of infecteds in the population is steady. When the epidemic is expanding, *R_e_* becomes larger than 1. On the other hand if *R_e_* becomes less than 1, the epidemic is declining. When 0 < *R_e_* < 1, the convergence of the epidemic is achieved by the recovery of patients. The measure which makes *R_e_* < 1 must be introduced by each government suitable to the country to accelerate the convergence.

## Data Availability

Ministry of Health, Labour and Welfare, Japan,
"COVID-19 cases reported on Apr. 29, 2020, Japanese)"

https://www.mhlw.go.jp/content/10906000/000626140.pdf

## Acknowledgments

I would like to thank Drs. M. Matsushita, M. Sano, Y. Yamazaki and R. Fujie for valuable discussion. My deepest appreciation goes to my family who helped me in preparation of this manuscript.

